# Agility and sustainability: A qualitative evaluation of COVID-19 Non-pharmaceutical Interventions (NPIs) in the UK logistics sector

**DOI:** 10.1101/2022.01.28.22270013

**Authors:** Hua Wei, Sarah Daniels, Carl A. Whitfield, Yang Han, David W. Denning, Ian Hall, Martyn Regan, Arpana Verma, Martie van Tongeren

**Affiliations:** Division of Population Health, Health Services Research & Primary Care, School of Health Sciences, University of Manchester, Manchester, England; Department of Mathematics, University of Manchester, Manchester, England; Division of Infection, Immunity & Respiratory Medicine, School of Biological Sciences, University of Manchester, Manchester, England; Manchester Academic Health Science Centre, University of Manchester, Manchester, England; Public Health, Advice, Guidance and Expertise, UK Health Security Agency, London, England; National COVID-19 Response Centre, UK Health Security Agency, London, England

**Keywords:** COVID-19, rapid response, logistics sector, delivery workers, non-pharmaceutical interventions

## Abstract

**Background:** The emergence of SARS-CoV-2 triggered a chain of public health responses that radically changed our way of living and working. Non-healthcare sectors, such as the logistics sector, play a key role in such responses. This research aims to qualitatively evaluate the non-pharmaceutical interventions (NPIs) implemented in the UK logistics sector during the COVID-19 pandemic.

**Methods:** We conducted nine semi-structured interviews in July-August 2020 and May-June 2021. In total 11 interviewees represented six companies occupying a range of positions in the UK’s logistics sector, including takeaway food delivery, large and small goods delivery and home appliance installation, and logistics technology providers. Inductive thematic analysis was completed using NVivo12 to generate emerging themes and subthemes. Themes/subthemes relevant to interventions were mapped deductively onto an adapted Hierarchy of Control (HoC) framework, focusing on delivery workers. Themes/subthemes relevant to the process of implementation were analyzed to understand the barriers and facilitators of rapid responses.

**Results:** HoC analysis suggests the sector has implemented a wide range of risk mitigation measures, with each company developing their own portfolio of measures. Contact-free delivery was the most commonly implemented measure and perceived effective. In addition, a broad range of measures were implemented, including social distancing, internal contact tracing, communication and collaboration with other key stakeholders of the sector. Process evaluation identified facilitators of rapid responses including capacity to develop interventions internally, localized government support, overwhelming external mandates, effective communication, leadership support and financial support for self-isolation, while barriers included unclear government guidance, shortage of testing capacity and supply, high costs and diversified language and cultural backgrounds. Main sustainability issues included compliance fatigue, and the possible mental health impacts of a prolonged rapid response.

**Conclusions:** This research identified drivers and obstacles of rapid implementation of NPIs in response to a respiratory infection pandemic. Existing implementation process models do not consider speed to respond and the absence or lack of guidance in emergency situations such as the COVID-19. We recommend the development of a rapid response model to inform the design of effective and sustainable infection prevention and control policies and to focus future research priorities.

**Contributions to the field:**

- The study offered important insights into the process of the UK logistics sector’s response to the COVID-19 pandemic.
- The Hierarchy of Control (HoC) framework was adapted for the evaluation of a collection of non-pharmaceutical interventions (NPIs) implemented in a non-healthcare sector.
- Thematic analysis of qualitative data generated themes that were relevant to the process of rapid implementation of NPIs during a public health emergency.
- Barriers, facilitators and sustainability issues of the sector’s rapid response to the COVID-19 pandemic have been highlighted to inform future research on implementation strategies.

## 1 Introduction

The novel severe acute respiratory syndrome coronavirus 2 (SARS-CoV-2) virus shocked the world in the last few days of 2019 and we still very much live in this Coronavirus disease 2019 (COVID-19) pandemic at the time of writing. In the UK, the logistics sector worked together to keep the workers and customers safe and increased capacity to cope with the sustained high level of demands. The sector employs and contracts a large number of workers to deliver a wide range products and goods to private and commercial addresses; many of them are self-employed. They could face both health and financial risks over a pandemic (1), and contribute to community transmissions (2-4). An analyses of COVID-19 mortality in England showed that, similar to other essential workers, van drivers had an increased risk of death from COVID-19, compared to non-essential workers (5). It is therefore important to introduce risk mitigation measures (RMMs) within this sector. Non-pharmaceutical interventions (NPIs) are often significant investments that require well-coordinated actions by multiple stakeholders across organizations and society (6, 7). To cope with imminent threats, such as a novel disease pandemic, interventions must be deployed rapidly to ensure behavioral and mindset changes occurring within a short time frame. In the case of COVID-19, mathematical models suggested that restrictive measures to reduce social mixing could reduce virus transmission and must take effect in a matter of days in order to save lives (8-10). While research about the health systems’ response to public health emergencies has provided good quality evidence (11, 12), similar evidence on the contribution of control measures in non-healthcare sectors, such as the logistics sector, to control work-related transmission is so far lacking (13-15). Hence, it is imperative to learn more about what RMMs were implemented by the UK logistics companies, the barriers and facilitators of implementation and whether the control measures are sustainable in the long-term. The aim of this study was to answer these questions through interviews that explored the company representatives’ opinions and experiences.

## 2 Material and Methods

We have generally followed the Consolidated Criteria for Reporting Qualitative Studies (COREQ) to report the methods and findings (16). A checklist can be found in Additional file 1.

### 2.1 Data collection

We recruited companies through a variety of approaches, such as direct contact, approaching trade and industry associations, via personal and professional networks and a social media campaign on LinkedIn. All recruitment activities were carried out using phones, emails or online facilities. We completed nine semi-structured interviews with six companies between July and August of 2020 (Round 1) and May and June of 2021 (Round 2), with three companies interviewed twice. Each of the interviews lasted between 60 and 90 minutes. There were in total 11 participants as four companies had two or three representatives.

All participants received a study scope and Participant Information Sheet and gave verbal consent before the interviews began. We used the Zoom teleconferencing facility to audio record the interviews. Three trained postdoctoral researchers (HW, SD, CW) carried out all the interviews, with attendance by other members of the study team. Interview schedules were developed in advance, with open ended questions which included inquiries on the type of RMMs implemented, facilitators and barriers of implementation, recommendations for possible future pandemics and potential health impacts of coping with a long pandemic. The interview schedules for both round 1 and 2 are available in Additional file 2. A summary report was emailed to each participating company for comments and corrections. One company returned written comments and another discussed feedback with us over Zoom.

### 2.2 Data analysis

HW, SD and CW edited and anonymized the auto-transcripts generated by Zoom. One company supplied a detailed list of events from February 2020 to July 2020, which was also analyzed. Thematic analysis was carried out using NVivo12 software following the latent approach (17, 18). HW and SD studied the transcripts and events list and identified emerging themes and sub-themes, which were developed into new codes to be fitted within the codebook.. Thereafter the themes and subthemes relevant to RMMs for delivery workers were deductively matched, if appropriately, with the levels of the Hierarchy of Control (HoC) (19, 20). HoC ranks preventative measures according to their expected level of protectiveness against one particular hazard, moving from the most protective measures that eliminate the hazard completely from the work environment, down to personal protective equipment (PPE), the last layer of protection for workers (see Figure 1). The mapping exercise was reviewed and discussed extensively within the team and with experts from the Health and Safety Executive (HSE) and Public Health England (PHE, now known as UK Health Security

**Figure 1.**
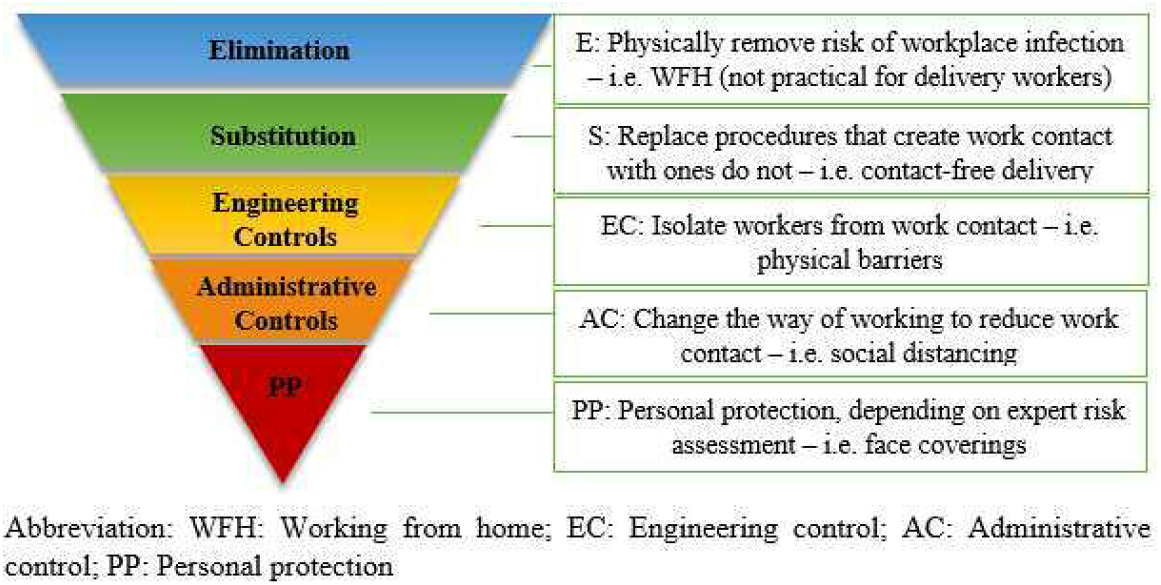
HoC: COVID-19 – Delivery workers (adapted from HSE websites^i^) The HoC analysis focused on the delivery workers who would collect deliveries from a workplace (i.e. warehouses or depots) and deliver them to customer premises, using a certain type of vehicle. For large items, they might also enter customer premises in order to drop the deliveries to a designated room (Room of Choice) or to complete the installation.

Agency). Codebooks were developed separately for the two rounds of interviews to allow for changes that occurred over the course of the pandemic. Based on the agreed codebooks, HW and SD then coded the transcripts independently, with coding results merged to assess inter-coder reliability. The percentage of agreement between the two coders was very high (>90%) and the average Kappa coefficient was 0.61 for the first round and 0.51 for the second round (0.41–0.75 is considered fair to good (21)). Individual codes that showed higher discrepancy were discussed and consensus was reached.

## 3 Results

### 3.1 Characteristics of participating companies

Participants represented one takeaway food delivery platform, four logistics companies that delivered large and small items and one technology provider for food and grocery chain stores i.e. supermarkets and restaurant chains. Most of the representatives that we recruited were directly involved in the day-to-day running of the logistics business. However, for grocery store deliveries, we only managed to recruit a technology developer that served the food and grocery chains. All the delivery companies were large employers (500+) except the technology developer. The roles of the participants covered a range of functions in the companies, including health and safety, operation, operational support, communication, marketing and external affairs. Delivery of large items was normally fulfilled by two-person teams, while parcel and takeaway food deliveries were fulfilled by lone drivers or bicycle riders. Of the five delivery companies, delivery workers were engaged as self-employed in four, with one large items delivery company employing drivers directly.

### 3.2 Interventions -- HoC analysis

HoC analysis focuses on the interventions. A wide range of RMMs were designed and implemented by the interviewed companies. Through the pandemic, they continued to do so to tackle newer challenges, such as the emergence of new variants, risks of increased transmission during the winter season, and adapting to new government measures, such as mass testing and vaccination. HoC analysis excluded the technology provider as they were not directly involved in delivery work. We summarized the RMMs that were discussed in the interviews in Table 1. Food 1 refers to the takeaway platform, Parcel 1 and 2 refer to the two parcel delivery companies, and Large 1 and 2 refer to the two large items delivery companies. Food 1 engages couriers using an app and does not operate any physical sites, while the other four companies do, of which, Large 1 and 2 also provide company vehicles.

**Table 1.** HoC analysis – COVID-19 RMMs implemented by the logistics companies for delivery workers.

| HoC/Measures | Food 1 | Parcel 1 | Parcel 2 | Large 1 | Large 2 |
| --- | --- | --- | --- | --- | --- |
| **1. Elimination: Physically remove risk of workplace infection** |  |  |  |  |  |
| None practical |  |  |  |  |  |
| **2. Substitution: Replace work procedures that create work contact with ones that do not** |  |  |  |  |  |
| Contact-free delivery | + | + | + | + | + |
| **3. Engineering Controls: Isolate workers from work contact** |  |  |  |  |  |
| Establish exclusion zones |  |  |  | + |  |
| Extra car hiring |  |  |  | Discussed but not adopted | + March-June 2020<sup>a</sup> |
| Install physical barriers |  | + | + | + | + |
| Re-layout workplace |  | + | + | + | + |
| Restricted or discontinued services |  | + Temporarily suspended customer collection | + Temporarily suspended customer collection | + Installation service suspended March-May 2020 | + RoC<sup>b</sup> suspended March-May 2020; Initially failed deliveries if customers reported symptomatic or self-isolating |
| Ventilation in buildings |  | Believed lack of airflow in winter was a cause of outbreaks | Deemed sufficient | Deemed sufficient | + Open windows |
| **4. Administrative controls: Change the way of working to reduce work contact** |  |  |  |  |  |
| Pairs and bubbles (staff cohorts) |  | + | + | + | + |
| Social distancing |  | + | + | + | + |
| Self-isolation (if symptomatic, tested positive or close contact) | + | + | + | + | + |
| Staggered working |  |  | + | + | + |
| Ventilation in shared vehicles |  |  |  | + Open windows | + Instructed windows 1/3 down and recirculation turned off |
| Hygiene measures | + | + | + | + | + |
| Information Instruction & Training (IIT) | + | + | + | + | + |
| Working with industry and authorities | + | + | + | + | + |
| Mental health support |  | + |  | + | + |
| Compliance behaviour monitoring | + | + | + | + | + |
| Workplace contact tracing |  | + | + | + | + |
| Workplace infection monitoring |  | + | + | + | + |
| Workplace testing |  | + Deployed 3rd party testing at sites had outbreaks | Had concerns about regular workplace LFD<sup>c</sup> testing | Had concerns about regular workplace LFD testing | Some sites used LFD for warehouse staff |
| Disciplinary action |  | + |  | + |  |
| **5. Personal protection: Protect workers with certain equipment, depending on expert risk assessment<sup>d</sup>** |  |  |  |  |  |
| Face coverings | + | + | + | + | + |
| Gloves | + | + |  | + |  |
| “+” indicates the measure was reported as implemented. This table is not a complete list of RMMs implemented by the companies. When some of the measures were not ticked by certain companies, it meant that this measure was neither applicable to the company’s situation nor discussed during the interviews.<br>a. Time period was estimated by the interviewers during analysis. b. RoC: room of choice c. Lateral Flow Device d. Neither face coverings nor normal gloves were considered PPE. They were issued to prevent transmission rather than protecting workers from getting infected. |  |  |  |  |  |

No measures taken by the companies fell within the definition of Elimination. For example, working from home (WFH) would eliminate risk of infection from workplaces but is not practical for delivery workers. “Other staff (i.e. office workers) WFH” is treated as an administrative control (AC) measure as it would help reduce workplace contacts for delivery workers.

Contact-free delivery was considered a Substitution measure and the most practical in the context of home deliveries. All five companies named it as the most important measure to reduce contacts for delivery workers and introduced it from a very early stage of the pandemic. It was achieved by drivers doing doorstep drop-off with no signature required. Proof of delivery that previously required customers to sign a paper document or the handheld unit with a pen, a finger or a wand was replaced by taking a photo at the doorstep or signed by the driver’s colleague when it was two-person deliveries.

> *“The moment the UK went into lockdown and we moved to doorstep delivery only*.*”* [Large 1]
>
> *“As soon as lockdown was announced*… *we stopped (delivery to) room of choice as well*.*”*

[Large 2]

> *“So quite quickly we had to establish a way of how could we achieve that without actually getting someone to touch our equipment or interact with the driver*… *And the way we achieved it is we took a photograph*… *It was accepted very quickly that that was the new form of signature*.*”* [Parcel 1]
>
> *“As well as asking drivers to knock on the door and then step back, we’ve also stopped getting signatures*.*”* [Parcel 2]
>
> *“We rolled out contact-free delivery across our entire network*… *so everybody was doing contract free delivery*.*”* [Food 1]

In terms of engineering controls (EC) measures, four of the companies that operated physical sites had installed physical barriers, changed workplace layout and restricted or suspended some services. One of them reported they erected temporary facilities such as portaloos and resting areas for visitors and third-party drivers. Companies took different measures to minimize contact for two-person deliveries. For example, Large 2 hired additional cars for the second delivery personnel so that the two-person team did not need to share the vehicle.

They stopped the measure following publication of government guidance on sharing vehicles at work in June 2020. Large 1 suspended installation service immediately following the first lockdown and resumed it when the government guidance about working in customers’ homes was introduced and they were able to establish safe work practice.

Most control measures reported were at the AC level. All five companies reported implementing self-isolation (if symptomatic, tested positive or close contact), hygiene measures, Information Instruction & Training (IIT), working with industry and authorities and compliance & data monitoring. Measures relevant to IIT or communication were described the most frequently by the participants. All participants discussed how they communicated the guidance, instructions and the changes to their employed or self-employed drivers, employees and customers throughout the period. These included daily or weekly bulletins, virtual Town Hall meetings, emails, phone texts, messaging platforms such as Yammer, YouTube channels, face-to-face briefings (if workspace allowed social distancing) and educational phone calls when issues arose. They monitored COVID compliance by collecting information via staff surveys, customer feedback, observational monitoring by dedicated staff or CCTV and site audits. All of them reported thorough promotion of hand wash and enhanced cleaning routines.

All four companies that operated from distribution centers implemented pairs and bubbles, social distancing, workplace contact tracing and workplace infection monitoring. Pairing refers to fixing each two-person delivery team permanently. Before the pandemic these pairs would change every day or in some cases multiple times per day. Drivers and warehouse staff would be grouped by location to establish working group bubbles, with no rotation between sites. The key was to keep the same teams together as much as possible to reduce the number of contacts, and to make workplace contact tracing more effective. When a case was confirmed, the workers who had been in close contact with the infected individual would be notified immediately to go into self-isolation. The other AC measures reported included staggered working where breaks and beginning of shifts were staggered at intervals, i.e. 15 minutes to minimize contact. All of the interviewed companies demonstrated a strong capacity in workplace infection rate monitoring, especially in the second round of interviews. Four of them stated infection rates in the workforce merely reflected community infection rates, indicating limited workplace transmission. One reported they had outbreaks within workplaces when the Alpha variant emerged in winter 2020. They then immediately deployed third-party testing facilities to test the entire workforce at those sites.

For personal protection and personal hygiene, participants reported they provided drivers with face coverings, gloves and hand sanitizers.

### 3.3 Implementation – process evaluation

In this section, we investigate the process of implementation and identify barriers, facilitators and issues that might affect sustainability. The process had prominent features, such as the speed to action, the overwhelming external pressure, improvised interventions, ad hoc approach, a fast-evolving situation and steep learning curves for all stakeholders. Based on the emerging themes of our thematic analysis, we summarized 15 key characteristics of rapid responses that can be categorized into five domains, with relevant barriers and facilitators identified in Table 2.

**Table 2.** Rapid response process: COVID-19 – Logistics sector, adapted for evaluating a range of RMMs.

| **Domain** | **Key characteristics** | **Illustrations** | **Barriers** | **Facilitators** |
| --- | --- | --- | --- | --- |
| Intervention characteristics | Source of interventions | Whether the interventions are perceived as externally or internally developed | Unclear or changing government guidance | Capacity to develop interventions internally |
|  | Evidence Strength & Quality | Whether data are collected about the effectiveness of the interventions and how the quality and validity of evidence are perceived | Shortage of testing capacity and supply |  |
|  | Costs | Direct costs of the interventions and costs associated with implementing the interventions including investment, supply, and opportunity costs. | High direct and associated costs |  |
| External environment | Prioritization of safety | The extent to which workers and customers' safety are prioritized by the organization and other external actors, such as the government. |  |  |
|  | Collaborations | The degree to which an organization is collaborating with other external organizations |  | Localized government support |
|  | External pressure | External pressure to enact a rapid response, such as government mandates or peer pressure i.e., other organizations have already implemented interventions |  | Overwhelming external mandates |
| Organizational setting | Effective communications | How the effectiveness and quality of communications are perceived | Diversified language and cultural backgrounds. | Effective communication |
|  | Safety culture | Norms, values, and basic assumptions about safety in the organization. |  |  |
|  | Implementation climate | The internal tension for change and the extent to which use of the interventions will be rewarded, supported, and expected within the organization. |  | Financial support for self-isolation |
|  | Leadership commitment | Commitment and involvement of leaders and managers with the implementation |  | Leadership support |

| Domain | Key characteristics | Illustrations | Barriers | Facilitators |
| --- | --- | --- | --- | --- |
| Implementation process | Rapid response | The degree to which the interventions are rapidly developed and implemented without planning in advance |  |  |
|  | Full engagement | Engaging appropriate individuals in the implementation of the interventions through a combined strategy of social marketing, education, role modelling, training, and other similar activities. |  |  |
|  | Strong execution | Carrying out or accomplishing the implementation according to plan |  |  |
|  | Continuous reflecting & evaluating | Continuous risk assessment and learning, accompanied with regular quantitative and qualitative feedback about the progress and quality of implementation |  |  |
| Sustainability | Potential long-term effects | Any possible long-term effects when rapid responses have lasted longer than expected |  |  |

#### 3.3.1 Intervention characteristics

The themes that emerged in relation to interventions characteristics included source of interventions, strength & quality of evidence and costs.

The companies developed the interventions drawing from both external and internal sources. External sources were mostly government guidance such as social distancing, face covering and hand washing, which were relatively standardized. For companies that operate in multiple countries, signals from other countries also provided sources of intervention. For example, Parcel 1 mentioned they had secured a supply of facemasks (described as three-layer paper masks) for their UK workers, as colleagues from across the world recommended this as a preventative measure at the early stages of the pandemic. Internally developed measures generally followed the principal of minimizing contact but with customized characteristics. Contact-free delivery is an example of an internally developed intervention with slightly different features designed by each company. Both Parcel 1 and 2 used photographs to replace customer signatures, while Large 1 required no signature and Large 2 asked the driver’s “mate” (the other personnel in a two-person delivery team) to sign as a proof of delivery. Food 1 required no signature and strongly advised online payment. When cash payment was necessary, they then asked the money to be put into an envelope.

##### Barrier 1

Barriers to rapid development of interventions here appeared to be the lack of and changing government guidance.

##### Facilitator 1

The resourcefulness and capacity to design and develop interventions internally appeared to be a facilitator.

The companies reported how they actively collected data to monitor the effectiveness of communication and infection rates. They mentioned customer and staff surveys, monitoring message click rate and dwelling time, and monitoring infection and self-isolation rates.

Participants appeared to be more confident about the quality and validity of the evidence in round 2. During round 1, they generally reported a very low number of confirmed cases, while during round 2, participants provided more details about how they collected and analyzed data systematically. They were able to make clear statements about the perceived cause of the outbreaks. For example, Large 1 discussed how the Alpha variant, combined with lack of ventilation in the winter season, had a significant impact on transmission in the workplace. They were clear about timing, location and job roles that were the most affected. Parcel 2 showed to us over Zoom their COVID infection dashboard where data were systematically collected, analyzed and displayed for decision making.

##### Barrier 2

Limited testing capacity and shortage of supply at the beginning of the pandemic appeared to be major barriers. This capacity was visibly improved during the course of the pandemic as demonstrated by the round 2 interviews.

##### Barrier 3

NPIs implemented at speed appeared to be costly. The participants talked about direct and associated costs including investment, supply or equipment and the knock-on effect on efficiency. Interventions such as deploying more vehicles, providing equipment and furniture to allow office staff to WFH, and providing hand sanitizers and face coverings would obviously add to costs. Financial support, such as 14-day COVID sick leave pay for the self-employed and additional bonuses, were direct costs. There was also other investment such as communication systems, posters and markings, sanitary stations, physical barriers and alteration of workplaces. One participant mentioned that costs related to COVID-19 RMMs were in the range of hundreds of thousands of pounds every month at that time. However, participants were also in universal agreement that the companies had been performing well since the first lockdown as demand increased and sustained at high level, which might have helped to absorb the costs.

#### 3.3.2 External environment

Three themes emerged in relation to external environment, including prioritization of COVID safety, unprecedented collaboration within the industry and overwhelming external mandates to enact rapid responses.

The UK government imposed lockdown measures in March and November 2020 and January 2021 to stop non-essential contact and travel. Nevertheless, delivery of food and other essential supplies was recognized as essential work by the government. Hence worker and customer safety must be prioritized and the companies modified work procedures to reduce work contact, including suspension of services, such as installation or Room of Choice, and stopped procedures, such as signing on documents or equipment.

The level of collaboration within the industry was unprecedentedly high as reported by the participants. It included working with the sector including competitors, the government and international collaboration within the organizations.

##### Facilitator 2

Localized government support was a facilitator of the rapid response. Participants described working with the local police, Department for Environment, Food and Rural Affairs (DEFRA), HSE, PHE, National Health Service (NHS) and local authorities. When there was a high level of uncertainty, the companies appreciated the support from local authorities and local branches of HSE, PHE and unions. They would send their internal guidance and risk assessment to these bodies and obtain their opinions. The support was personalised to the companies, which then provided the companies with confidence to implement these measures.

Networking in this sector was strengthened especially at the beginning of the pandemic. Participants spoke highly about the industry forum organised by DEFRA that occurred weekly and then bi-weekly. It was unprecedented as all the main competitors of the industry joined. Participants reported that they shared best practices with an open mind and worked together to contribute to the development of government guidance. Email groups were set up to facilitate exchange of ideas and questions.

##### Facilitator 3

In addition to the networked collaborative activities, the numerous government recommendations, guidelines and updates, and that COVID-19 dominated the media and the Internet for a substantial period of time, all created an overwhelming incentive for the companies to respond rapidly.

#### 3.3.3 Organizational setting

Four themes emerged in this domain, including effective communications, safety culture, facilitating implementation climate and leadership commitment for implementation.

##### Facilitator 4

Effective communications were emphasized by many participants as an important facilitator of rapid responses. They reported that communications were highly valued by the staff because the situation had been a fast-evolving one. Uncertainties and lack of specific guidance meant that workers needed that information to guide their everyday work. Every time new threats, guidance or public measures came out, such as regional lockdowns, new variants, mass testing or the vaccination program, it was important the company communicated guidance and recommendations that were specific to their work context.

##### Barrier 4

A number of participants reported that language and the complexity of the guidance could be a challenge as English is the second language for many workers within this sector. To tackle this issue, they simplified the language and added infographics to illustrate the meaning. A couple of participants mentioned the cultural background of the workforce could be a barrier to enforce social distancing as certain cultures tend to socialise more and workers of that background were likely to share transport to work or accommodations. This however could also be a facilitator in some circumstances as Large 1 explained how they paired “sons and dads, mums and dads” or drivers from the same households or family bubbles for two-person deliveries.

COVID-19 safety was discussed by the participants as a belief rather than something they reluctantly comply to. One participant articulated it particularly well.

> *“We have a culture in our leadership of putting safety first*… *we track our [COVID-19] numbers in [Large 2] but there’s no incentive. You know I’m not bonused, my performance isn’t measured on whether I achieve safety or not. We all do it because it’s the right thing to do*.*”* [Large 2]

The organizational climate for implementing interventions played a facilitating role. Key stakeholders felt the necessity to change in order to keep safe and contain the spread of the virus, as one of the participants described:

> *“The behaviour change, the couriers, the restaurants, the customers was helped by the fact that every single aspect of life has changed. So people [were] kind of shocked into it*.*”* [Food 1]

##### Facilitator 5

Three of the companies mentioned they provided financial support such as sick leave pay to support the self-employed drivers to take COVID-related self-isolation. It can facilitate adherence among delivery workers as many of them were self-employed and did not enjoy statutory sick pay. They also mentioned that they promoted intangible incentives such as customers’ appreciation messages and exemplar stories to be put on their websites and communication channels.

##### Facilitator 6

Key stakeholders’ commitment for implementation appeared high. Leadership engagement was evident in all the interviews. Two participants particularly emphasized the influence from the leadership team that keeping workers safe from COVID-19 infection was the right thing to do and would reward the business in the long-term. This is then linked to resources dedicated for implementation. It appeared that the companies allocated adequate resources timely to support the interventions.

#### 3.3.4 Implementation process

The implementation process can be characterized as an unplanned rapid response, full engagement, strong execution and continuous reflecting & evaluating.

“Rapid response” was a prominent feature emphasized by all of the participants. From early March 2020, the volume of home deliveries “went through the roof”. Participants mentioned figures such as:

> *“Our sales spiked*… *202% year on year compared to previous March”* [Large 1]
>
> *“Volume of orders have gone up, way up, absolutely unbelievable”* [Logistics technology provider]

In response, the sector moved rapidly to increase the capacity, while ensuring worker and customer safety. Changes and interventions were obviously not planned in advance. Supply chain networks are underpinned by technology that help streamline the service. The technology provider participant described the chaos experienced by food and grocery chains during the first lockdown. Restaurants, cafes and small retailers were closed and hence the volume of that part of the supply chain went down to zero whilst supermarkets suddenly faced much higher demands which caused blockages and bottlenecks in their network. “It completely destroyed that (food) supply chain”, the participant recalled. Nevertheless, their engineers rose to the challenge and developed solutions for the clients in just six days. The participant told us internally the grocery chains called it “the second Christmas” as they “turned on the Christmas protocols for everything” in a matter of days, whereas normally preparations for the Christmas peak would take a few months.

Other participants also passionately described the speed of implementation.

> “*We were able to react really quickly. And we were able to get, as I’ve said, sort of, PPE, standards, working from home, all of those things in really, really quickly. We even surprised ourselves*… *we really pride ourselves on how quickly*… *and we’ve done it really smoothly*.*”* [Parcel 1]
>
> *“And so lots and lots of shared facilities across all of our sites that we just had to change pretty much overnight and because we didn’t stop operate so real big challenges*.*”* [Large 2]

There were many more examples that described deployment of interventions in a very short timeframe such as overnight, within a week, or in just a few days.

As an unplanned response, continuous risk assessment combined with an experimental approach were essential. There were measures that were considered but not adopted or were on hold for future review. This can be an important feature for learning when facing emergencies caused by novel threats in the future. Participants discussed these measures and reasons for not adopting them.

> *“We explored offering our people tests, we decided not to do that because there was a lot of uncertainty. This was around May [2020] time. There was a lot of uncertainty about which test, availability of tests*… *We wrestled with the ethics of if we take a big batch of tests. Does that take away from the NHS and care homes?”* [Large 1]
>
> *“The key reason we didn’t do that [ordering facemasks in bulks] immediately was because we wanted to ensure that what we were ordering wouldn’t impact the NHS and care homes receiving it*.*”* [Food 1]

When mass testing became available later in the pandemic, it was not immediately adopted by the companies. Participants reasoned that regular lateral flow device testing could not be easily integrated into their daily operations. One participant expressed a strong view regarding the possible effect of workplace testing in undermining other existing measures.

> *“Workplace testing when you’re dealing with certain members of society actually has a detrimental effect in terms of following COVID secure guidelines that we’ve put in place. So what we felt was that by introducing workplace testing people felt that was a level of security that I didn’t agree with, and that if they felt that they tested negative, then they didn’t need to follow social distancing wear face coverings so*….*my view is quite strong on this is that actually lateral flow testing undermines a lot of the measures that we really need people to be focusing on*.*”* [Parcel 2]

#### 3.3.5 Sustainability

A rapid response mode may be effective in the short-term but can run into problems if it lasted longer than expected and hence introduce questions about sustainability.

As the pandemic continued into 2021, some workers developed compliance fatigue and this became a barrier to effective implementation. In round 2 interviews, we asked the participants whether they observed any relaxed attitudes towards the COVID measures.

Participants agreed that to some extent attitudes had relaxed and described how they took actions to mitigate this. They highlighted the need to maintain effective communications by providing a “permanent alert” or “constant reminder” to workers. Two participants mentioned they added extra monitoring, that is, sending out staff to walk around the workplaces and giving colleagues a reminder whenever they observed behaviours not meeting the standard.

In round 2, all participants stated that high volume of home deliveries continued even when lockdown was lifted. They told us that the industry was used to working on full speed during the Christmas peak that was normally from late October to the end of December. As mentioned earlier, the industry immediately switched on the Christmas protocol from March 2020 and this continued into 2021. Mental health impacts of sustained high workload were mentioned by many of the participants. Participants expressed concerns about overwork, burnout and presenteeism.

> *“I’ve got a massive concern about burnout, about mental health, and you know the issues that overwork create*… *the level of additional work has just continued*… *it’s not just the burnout because you can bring the extra people in, it’s the prolonged on and on and on and on and no light at the end of the tunnel*.*”* [Parcel 1]

For office workers, while some appreciated the time saved from commuting by WFH, not all have an appropriate work environment in their homes and some reported feeling isolated.

Participants also mentioned that the companies were surveying workers regarding to their mental wellbeing and trying to offer some support.

We have provided a schematic diagram to illustrate the important findings in Figure 2. It highlights the key characteristics of rapid responses, grouped into five domains (interventions characteristics, external environment, organizational setting, and sustainability), with the implemented NPIs matched with HoC at the center.

**Figure 2.**
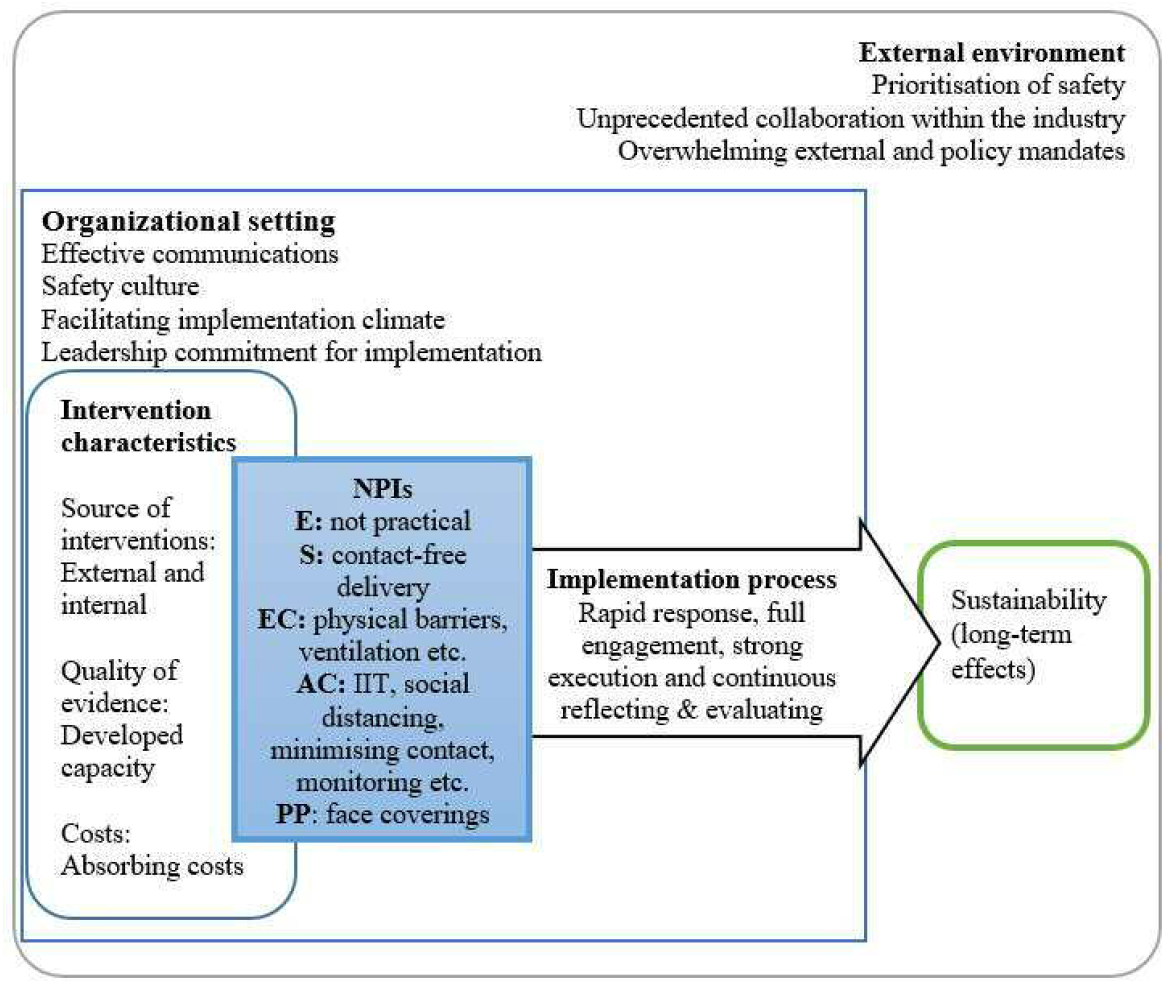
Qualitative evaluation of non-pharmaceutical interventions in non-healthcare sector: an example of the UK logistics sector during COVID-19 pandemic.

## 4 Discussion

This empirical research is responding to the call for knowledge and recommendations for preventive interventions to reduce the transmission of SARS-CoV-2. It offered a case analysis for the UK logistics sector, with an occupational focus on delivery workers.

The process of implementation had prominent features, such as the speed to action, the overwhelming external pressure, improvised interventions, and steep learning curves for all stakeholders. We scoped the literature to identify an appropriate theoretical model to inform the analysis. Multiple existing frameworks offered some useful insights, including the RE-AIM (Reach, Efficacy, Adoption, Implementation and Maintenance) (22, 23), CFIR (Consolidated Framework for Implementation Research) (24, 25), PRECEDE-PROCEED (26, 27) and other process evaluation models that generally included components such as recruitment, dose delivered, dose received, fidelity, satisfaction, maintenance and context (28). However, they generally assumed a systematically developed intervention program implemented with some extent of control, and none of them fully captured the characteristics of this sector’s response to COVID-19. It suggests the urgency of developing a rapid response model that can first, analyze a collection of NPIs implemented in occupational settings. When responding to a pandemic, NPIs are likely to be implemented simultaneously with many other measures and a single measure would not be sufficient (20). Second, the model should take into account the barriers and facilitators of rapid responses to a public health emergency (29).

In addition to the well-known COVID-19 NPIs, such as face coverings, hand washing and social distancing (14), our HoC analysis identified measures that were important to the delivery work setting, including contact-free delivery, fixed pairing, effective communications/IIT and sectoral collaboration. Contact-free delivery and fixed pairing (for two-person deliveries) were new measures improvised by this sector during this COVID-19 pandemic and became established practices as the participants told us. Working collaboratively with key stakeholders of the sector, including the competitors and local and state authorities was considered an important measure and a facilitator in outer setting (25).

We identified important barriers and facilitators to rapid responses. Financial support for self-isolation was considered a facilitator for delivery workers especially the self-employed, as a previous study found sick leave pay was associated with adherence to infection, prevention and control measures among healthcare workers (30). In addition, COVID-19 infection rates among delivery and warehousing workers from the developed and developing countries varied significantly. For example, in Canada, it was as low as 0% (31), whilst in Ecuador it was 15.2% (32). Although the sample of the two studies may not be directly comparable, it is possible that financial conditions served a social determinant of COVID-19 related health outcome (33). The sector’s capacity to design and develop interventions internally was also a key facilitator. As SARS-CoV-2 was a novel virus and the pandemic was fast-evolving, a response protocol or prevention guidance for the logistics sector was not available in the UK initially. Hence, internal knowledge and assessment was an important source of intervention development. Companies also used their judgement to decide not to adopt certain measures, such as workplace testing. This echoes the concern that people without COVID-19 self-isolating due to false-positive lateral flow test results could be a cost to the individual, their household, and their workplace (34). In addition, localized government support, effective communication and leadership support were considered facilitators. This is in line with findings from existing studies that evaluated the implementation of interventions programs (35, 36). Overwhelming external mandates was probably a prominent facilitator associated with the situation of a pandemic as few other health interventions received media attention like those for COVID-19.

Major barriers included unclear and changing government guidance, lack of testing capacity, shortage of facemasks, and diversified language and cultural backgrounds. Barriers associated with government guidance, testing capacity and supply of PPE mainly affected the rapid response at the early stages (37, 38). Language and cultural barriers were also identified by multiple intervention studies previously (36, 39). Carefully designed trainings were recommended, which were consistent with the measure took by the companies we interviewed. We identified compliance fatigue in the second interview round. Such behavioral changes reflected a response to adjustments in individuals’ risk assessment (40, 41), especially when the government announced their Roadmap to lift restrictions. Our participants suggested adding more behavior monitoring measures and reminders to maintain the level of alert. Participants mentioned the high costs associated with these NPIs but also believed such costs were compensated by increased volume. Going forward, a more systematic approach should evaluate such costs from health economics perspective.

The prolonged WFH measure and sustained high workload both add to work stress (42). It highlighted a key sustainability issue associated with the current approach to dealing with the pandemic. The concern is consistent with findings from studies that examined healthcare staff burnout during COVID-19 (43-45). It is not sustainable, and a more systematic approach and coherent sectoral strategy is urgently needed.

This paper is based on views expressed by those in managerial roles rather than the delivery drivers. We recognize that their views could differ significantly from the frontline workers’ perspective. For example, surveys among app-based drivers reported concerns of infection risks from interactions with the public and insufficient workplace protections such as access to personal protective equipment (PPE) (31, 46). Delivery workers in the French gig economy also expressed concerns of financial precarity and lack of union support (1).

Another potential limitation of this study is the small sample size and the size of the participated companies. The sector was extremely busy throughout the pandemic and our invitations were declined by the majority of companies we approached. We were not able to directly assess the effectiveness of the interventions, but the perceived effectiveness of the participants.

## 5 Conclusion

This qualitative study provides a rich source of contextualized data to evaluate rapid implementation of COVID-19 NPIs in the UK logistics sector. We assessed the interventions against an occupational health and safety standard and identified barriers, facilitators and sustainability issues in the process of a rapid response. In conclusion, the UK’s logistics sector rose to the challenge and rapidly developed and implemented a wide range of RMMs in a fast-evolving pandemic. They closely followed national and local guidelines available to them at the time and developed RMMs resourcefully when guidelines were lacking.

Elimination of the risk was not practical for the delivery workers and most control measures were considered administrative controls. Contact-free delivery was commonly implemented and considered effective. Participants were confident that the RMMs played an important role in reducing workplace transmission risk for delivery workers. Further research is now needed to design and evaluate models and tools to apply sustainable respiratory infection prevention and control measures across work settings, as well as taking into account the more vulnerable work and social groups.

## Supporting information

COREQ 32-item checklist

## Data Availability

All data produced in the present work are contained in the manuscript

## 6 Ethics statement

The project was reviewed and approved by the University Research Ethics Committee at University of Manchester, Ref: 2020-9787-15953. Consent to participation was verbally obtained before the commencement of the interviews.

## 7 Data availability statement

The datasets generated for this study are available on request to the corresponding author.

## 9 Author Contributions

Interview schedules and codebooks were developed by HW, SD, CW and MvT with extensive inputs from the rest of the team and collaborators at HSE and PHE. HW, SD and CW carried out the interviews and completed the analysis. HW and SD drafted the paper. MvT, YH, DD, IH, MR and AV commented and edited each version of the manuscript. All the authors reviewed and approved the final version.

## 11 Acknowledgments

The authors would like to thank the project’s advisory group that consist of Catherine Noakes, Chris Armitage, Sheena Johnson, Jeanette Edwards, Barbara Hockey, Nina Day, Nick Gent and Thomas House, for their advice that helped refine the aims and objectives of this article. We would also like to thank Helen Beers and Peter Baldwin from HSE for their advice on the business engagement, thematic analysis and feedback to companies.

