## Supplementary material for "Agility and sustainability: A qualitative evaluation of COVID-19 Non-pharmaceutical Interventions (NPIs) in the UK logistics sector": COREQ 32-item checklist

### Additional file 1: Consolidated criteria for reporting qualitative studies (COREQ): 32-item checklist

Developed from:

Tong A, Sainsbury P, Craig J. Consolidated criteria for reporting qualitative research (COREQ): a 32-item checklist for interviews and focus groups. *International Journal for Quality in Health Care*. 2007. Volume 19, Number 6: pp. 349 – 357

| **No. Item** | **Guide questions/description** | **Reported on Page #** |
| --- | --- | --- |
| **Domain 1: Research team and reﬂexivity** |  |  |
| *Personal Characteristics* |  |  |
| 1. Interviewer/facilitator | Which author/s conducted the interview or focus group? | Page 3 |
| 2. Credentials | What were the researcher’s credentials? E.g. PhD, MD | Page 3 |
| 3. Occupation | What was their occupation at the time of the study? | Page 3 |
| 4. Gender | Was the researcher male or female? | Not necessary to report |
| 5. Experience and training | What experience or training did the researcher have? | Page 3 |
| *Relationship with participants* |  |  |
| 6. Relationship established | Was a relationship established prior to study commencement? | Page 3  . |
| 7. Participant knowledge of the interviewer | What did the participants know about the researcher? e.g. personal goals, reasons for doing the research | Page 3 |
| 8. Interviewer characteristics | What characteristics were reported about the interviewer/facilitator? e.g. Bias, assumptions, reasons and interests in the research topic | Page 3 |

| **Domain 2: study design** |  |  |
| --- | --- | --- |
| *Theoretical framework* |  |  |
| 9. Methodological orientation and Theory | What methodological orientation was stated to underpin the study? e.g. grounded theory, discourse analysis, ethnography, phenomenology, content analysis | Page 3 & 4 |
| *Participant selection* |  |  |
| 10. Sampling | How were participants selected? e.g. purposive, convenience, consecutive, snowball | Page 3 |
| 11. Method of approach | How were participants approached? e.g. face-to-face, telephone, mail, email | Page 3 |
| 12. Sample size | How many participants were in the study? | Page 3 |
| 13. Non-participation | How many people refused to participate or dropped out? Reasons? | Page 13 |
| *Setting* |  |  |
| 14. Setting of data collection | Where was the data collected? e.g. home, clinic, workplace | Page 3  . |
| 15. Presence of non-participants | Was anyone else present besides the participants and researchers? | None – page 3 |
| 16. Description of sample | What are the important characteristics of the sample? e.g. demographic data, date | Page 3 & 4 |
| *Data collection* |  |  |
| 17. Interview guide | Were questions, prompts, guides provided by the authors? Was it pilot tested? | Additional file and page 3 |
| 18. Repeat interviews | Were repeat interviews carried out? If yes, how many? | Yes, three -- Page 3 |
| 19. Audio/visual recording | Did the research use audio or visual recording to collect the data? | Page 3 |
| 20. Field notes | Were ﬁeld notes made during and/or after the interview or focus group? | No – page 3 |
| 21. Duration | What was the duration of the interviews or focus group? | Page 3 |
| 22. Data saturation | Was data saturation discussed? | Page 3 & 4 |
| 23. Transcripts returned | Were transcripts returned to participants for comment and/or correction? | Page 3 |
| **Domain 3: analysis and ﬁndings** |  |  |
| *Data analysis* |  |  |
| 24. Number of data coders | How many data coders coded the data? | Page 3 |
| 25. Description of the coding tree | Did authors provide a description of the coding tree? | Page 4 |
| 26. Derivation of themes | Were themes identiﬁed in advance or derived from the data? | Page 3 & 4 |
| 27. Software | What software, if applicable, was used to manage the data? | Page 3 |
| 28. Participant checking | Did participants provide feedback on the ﬁndings? | Page 3 |
| *Reporting* |  |  |
| 29. Quotations presented | Were participant quotations presented to illustrate the themes/ﬁndings? Was each quotation identiﬁed? e.g. participant number | Yes. Yes. Page 5, 8- 11 |
| 30. Data and ﬁndings consistent | Was there consistency between the data presented and the ﬁndings? | Yes,  Page 4 to 11 |
| 31. Clarity of major themes | Were major themes clearly presented in the ﬁndings? | Yes,  page 4 to 11 |
| 32. Clarity of minor themes | Is there a description of diverse cases or discussion of minor themes? | Yes,  page 4 to 11 |
